# Effects of childhood and adult height on later life cardiovascular disease risk estimated through Mendelian randomization

**DOI:** 10.1101/2023.10.12.23296922

**Authors:** Tom G Richardson, Helena Urquijo, Laurence J Howe, Gareth Hawkes, Timothy M Frayling, George Davey Smith

## Abstract

**Background:** Taller individuals are at elevated and protected risk of various cardiovascular disease endpoints. Whether this is due to a direct consequence of their height during childhood, a long-term effect of remaining tall throughout the lifecourse, or confounding by other factors, is unknown.

**Methods:** We sought to address this by harnessing human genetic data to separate the independent effects of childhood and adulthood height using an approach known as lifecourse Mendelian randomization (MR). We analysed 5 cardiovascular disease endpoints (coronary artery disease (CAD), stroke, peripheral arterial disease (PAD), atrial fibrillation (AF) and thoracic aortic aneurysm (TAA)) using findings from large-scale genome-wide consortia (n=184,305 to 1,030,836).

**Results:** Protective effects of taller childhood height on risk of later life CAD (OR=0.78 per change in height category, 95% CI=0.70 to 0.86, P=4×10 ^−10^) and stroke (OR=0.93, 95% CI=0.86 to 1.00, P=0.03) were found using a univariable model, although evidence of these effects attenuated in a multivariable setting upon accounting for adulthood height. In contrast, direct effects of taller childhood height on increased risk of later life AF (OR=1.61, 95% CI=1.42 to 1.79, P=5×10 ^−7^) and TAA (OR=1.55, 95% CI=1.16 to 1.95, P=0.03) were found even after accounting for adulthood height in the multivariable model.

**Conclusions:** The protective effect of childhood height on risk of CAD and stroke is largely attributed to the causal pathway involving adulthood height, which may therefore be explained by taller children typically becoming taller individuals in later life. Conversely, the independent effect of childhood height on increased risk of AF and TAA may point towards developmental mechanisms in early life which confer a lifelong risk on these disease outcomes.

## Introduction

Height in adulthood has been associated with overall and cause-specific morbidity and mortality risk for over a century (1, 2). Taller height is generally associated with protective effects on overall health status and mortality, as well as lower incidence of most forms of cardiorespiratory disease (3). In contrast, taller height is associated with increased risk of several non-smoking related cancer sites (2, 3). In addition to studies of adult height, taller prepubertal childhood height has been related to the risk of cardiovascular outcomes (4).

Several mechanisms have been postulated to account for the associations of height and cardiovascular disease. These include mechanical effects (e.g. greater height leading to wider artery bore, or better lung function), linear growth programming later cardiovascular risk factors, confounding by early life environmental factors that increase both the risk of cardiovascular disease and reduced final height due to factors such as infections, nutritional deficiency and illnesses, and/or genetic pleiotropy. Additionally, genetic variation may contribute towards these mechanisms, including influences on Marfan syndrome and *formes frustes*, for which increased risk of thoracic aortic aneurysm is a distinctive feature (3, 5).

Mendelian randomization (MR)(6–9) - an approach using germline genetic variants as proxies for exposures and phenotypes putatively influencing disease risk, which mitigates some of the interpretive issues in naïve observational epidemiological studies – has been applied to investigate the causal processes involved (10–12). These studies provide support for some hypotheses regarding mechanisms underlying the link between adulthood height and health.

Observational evidence from the literature regarding childhood height and later life health is limited compared to that of adulthood height. Furthermore, MR studies concerning this issue are lacking. We have demonstrated that it is possible to separate the effects of prepubertal childhood adiposity and later life adiposity using MR (13–17), and here we extend this approach to examine the influence of childhood and adulthood height on cardiovascular disease endpoints.

## Methods

### Data resources

#### The UK Biobank study

Data from the UK Biobank study (UKB)(18) was used to identify genetic variants robustly associated with childhood and adult height. This protocol to derive genetic instruments was applied previously for a similar pair of variables based on childhood and adult body size (13). For childhood height, UKB participants were asked during their initial clinical assessment ‘when you were 10 years old, compared to the average would you describe yourself as shorter, taller or about average?’. This data was coded as an ordered categorical variable (i.e. 0=shorter, 1=about average and 2=taller). At the same clinic visit participants also had their standing height measured in centimetres (cm) using a Seca 202 device. This continuous trait was categorised into 3 groups based on the same proportions as the childhood height variable (i.e. ‘shorter’, ‘about average’ and ‘taller’). We did this to harmonise our childhood and adult measures and make them as comparable as possible for downstream analyses. Effect estimates for both childhood and adult height in this study can therefore be interpreted as odds conferred per additive change in height category.

Details on UK Biobank genotyping and imputation used in this study have been described previously (19). In brief, 12,370,749 genetic variants across the human genome passed quality control filtering. Analyses were restricted to individuals of predicted European descent (based on K means clustering (K=4)) after standard exclusions such as withdrawn consent, mismatch between genetic and reported sex, and putative sex chromosome aneuploidy. In total, there were 454,023 individuals with both childhood and adult measures of height as well as genetic data who were eligible for GWAS analyses.

#### Cardiovascular disease outcomes

We used summary statistics derived from large-scale consortia to estimate effects on 5 cardiovascular disease outcomes whose aetiology has been previously linked with height. These included coronary artery disease (CAD) (60,801 cases and 123,504 controls) (20), peripheral artery disease (PAD) (12,086 cases and 449,548 controls) (21), stroke (40,585 cases and 406,111 controls) (22) and its subtypes, atrial fibrillation (60,620 cases and 970,216 controls) (23) and thoracic aortic aneurysm (7,321 cases and 317,899 controls from the FinnGen study) (24). A summary of the study characteristics for these outcomes can be found in **Supplementary Table 1**.

### Statistical analysis

#### Identifying genetic instruments for childhood and adult height

GWAS analyses of childhood and adult height in UKB were undertaken using the BOLT-LMM software which uses a genetic relationship matrix between samples to account for relatedness and population stratification (25). Analyses were adjusted for age, sex and a binary variable to indicate which genotyping chip was used to obtain genetic data in participants. GWAS were undertaken firstly in all 454,023 participants in UKB who had both measures of childhood and adult height. We undertook linkage disequilibrium (LD) clumping on each set of GWAS results in turn to identifying independent genetic variants robustly associated with height (based on P<5×10 ^−08^). A reference panel of 10,000 randomly selected Europeans from UKB was used to calculate LD between variants so that all instruments were independent of each other based on r ^2^<0.001 (26).

#### Validation of genetic instruments

We conducted several validation analyses of our genetic instruments to demonstrate that they were able to reliably separate the effects of childhood and adult height. This was particularly warranted due to our childhood measure being derived using questionnaire recall data which could potentially lead to bias without appropriate validation using external datasets. This consisted of evaluations of how well our genetic predictors were capable of separating height measured during childhood and adult hood timepoi nts in two UK birth cohorts: the Avon Longitudinal Study of Parents and Children (ALSPAC) (27, 28) and 1958 National Child Development Study (1958 British Birth Cohort) (29). Additionally, we applied LD score regression to estimate genetic correlations between our genetic instrument sets and results from previously conducted GWAS of measured childhood (30) and adult height (31). We also repeated our GWAS analyses on 38,807 siblings with available phenotype data from the UKB study to allow validation of our derived instruments using within-sibship models, which are considered to be more robust to sources of bias induced by factors such as assortative mating and dynastic effects (32).

Lastly, we compared the correlation of childhood and adult height measures between spouse pairs to further investigate evidence of assortative mating for these traits as described previously (33). Full details on these validation analyses can be found in **Supplementary Note 1.**

#### Two-sample Mendelian randomization

Univariable MR analyses were firstly conducted to estimate the total effects of childhood and adult height (**Figure 1a**) on each of the cardiovascular endpoints in turn. This was initially assessed using the inverse variance weighted (IVW) method, which uses all SNP-outcome estimates regressed on those for the SNP-exposure associations to provide an overall weighted estimate of the causal effect based on the inverse of the square of the standard error for the SNP-outcome association(34). In the form of sensitivity analysis, we applied the weighted mode, weighted median and MR-Egger methods to evaluate the robustness of IVW estimates to horizontal pleiotropy (35–37).

**Figure 1:**
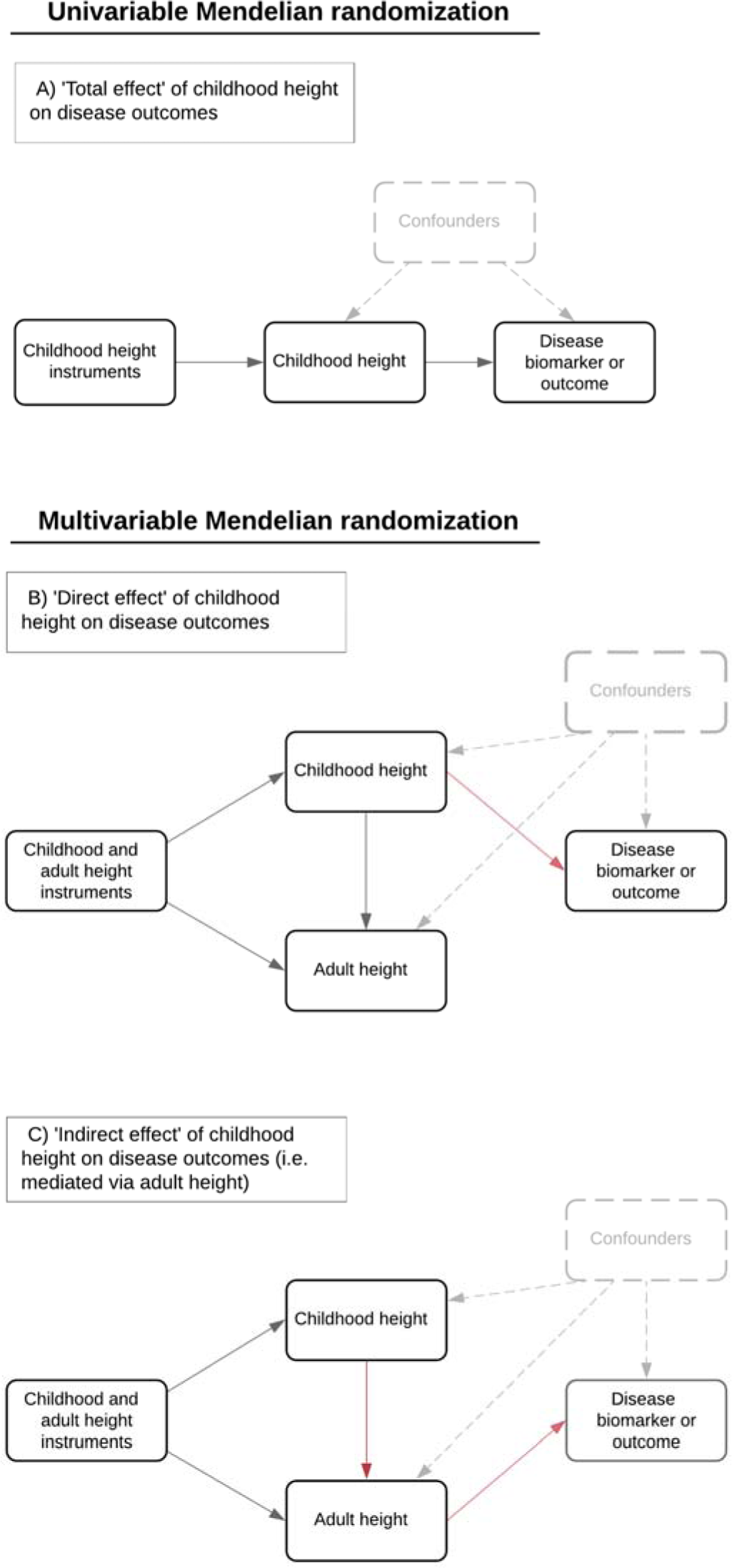
Directed acyclic graphs outlining analyses in this study to estimate A) ‘total effects’ of height on disease outcomes and biomarkers using univariable Mendelian randomization (MR) B) multivariable MR analyses to estimate ‘direct effects’ of childhood height on outcomes and C) undertaking multivariable MR to estimate ‘indirect effects’ on outcomes which are mediated via adult height.

We next applied two-sample multivariable MR to simultaneously evaluate the effects of childhood and adult height on these cardiovascular outcomes (38, 39). This allowed us to estimate the ‘direct effect’ of childhood height on outcomes (i.e. the effect independent of adult height (**Figure 1b**)) as well as the ‘indirect effect’ (i.e. the effect that childhood height has on outcomes along the causal pathway which is mediated by adult height (**Figure 1c**)). As a sensitivity analysis, we repeated analyses on thoracic aortic aneurysm after excluding instruments within genomic regions responsible for encoding the fibrillin family of proteins (i.e. *FBN1*, *FBN2* and *FBN3* +/-1Mb) given that mutations in these genes are known to cause Marfan’s syndrome and increase height (5, 40). Additionally, we analysed natural hair colour as an outcome to evaluate potential bias due to population stratification as part of a negative control analysis (i.e. given that childhood/adult height cannot influence natural hair colour) (41).

All analyses were undertaken using R (version 3.5.1). MR and sensitivity analyses were undertaken using the ‘TwoSampleMR’ package. (42). ROC curves were generated using the ‘pROC’ package and all other plots were created using the ‘ggplot2’ package (43).

## Results

### Identification and validation of genetic scores for childhood and adult height

Our GWAS identified 840 and 1201 independent genetic variants in UKB participants that were robustly associated with childhood and adult height measures respectively (based on P<5×10 ^−08^) (**Supplementary Tables 2 and 3**). The estimated proportion of variance explained by our GWAS results were 7.7% and 12.9% for childhood and adult height respectively with a pairwise genetic correlation of rG=0.89 (95% CI=0.88 to 0.90).

Constructing these variants as genetic risk scores in the ALSPAC cohort found that the genetic score for childhood height was a stronger predictor of height at mean age 9.9 years (n=6,222) compared to its adult counterpart (0.685 vs 0.666 AUC for childhood and adult scores respectively). Conversely, the adult score was a stronger predictor of height compared with our derived childhood score (0.701 vs 0.732 AUC) using the adult measured height (mean age: 47.9 years) in the ALSPAC cohort (**Figure 2**). The childhood height score also explained more of the variance in height measured at age 10 years using data from the 1958 British Birth Cohort (r ^2^=0.122) compared to the adulthood height score (r ^2^=0.116). Conversely, the adulthood height score explained more of the variance in height measured at age 44 years (r ^2^=0.206) compared to the childhood height score (r ^2^=0.163). The dominance of the childhood height score reverses by age 16 (see **Figures S1 & S2** in **Supplementary Note 1**).

**Figure 2:**
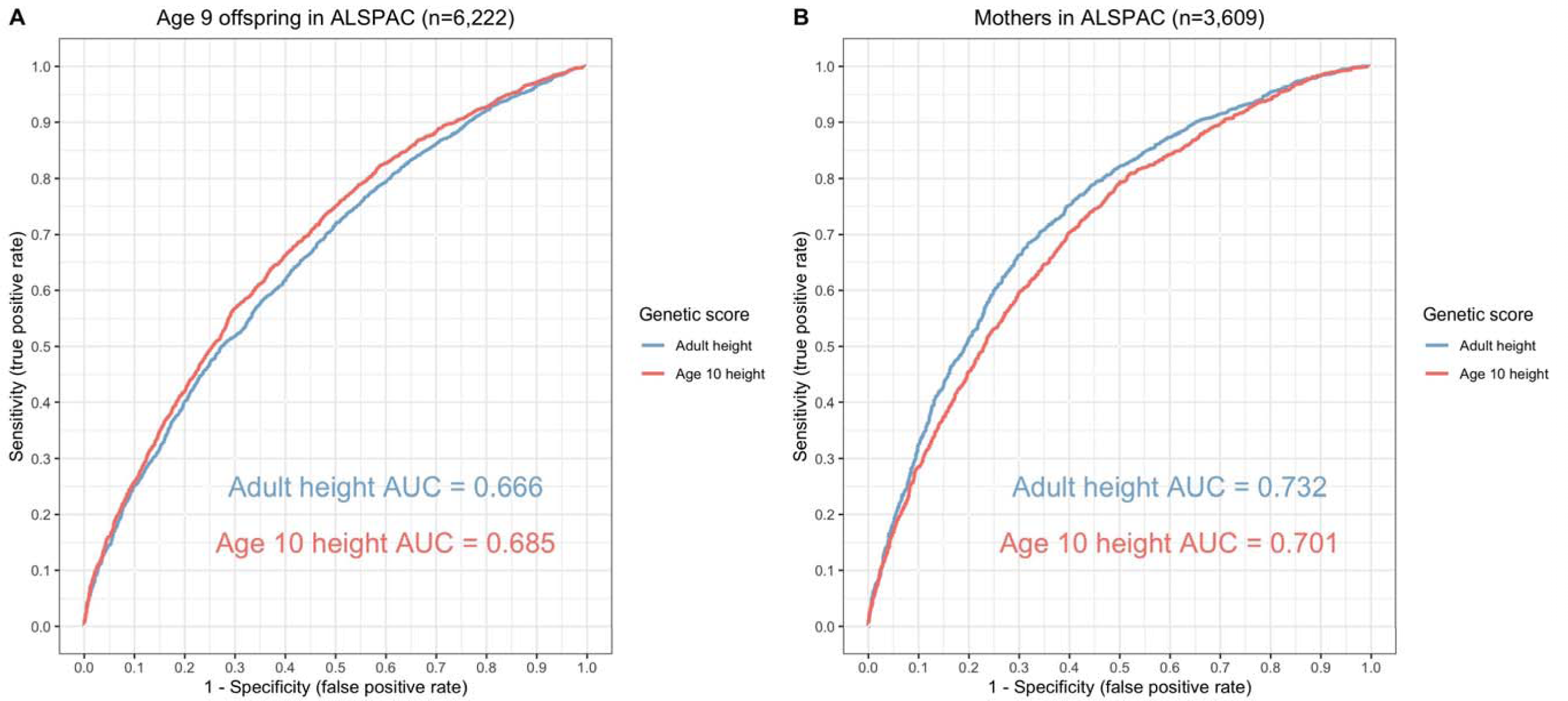
Validation Receiver operator characteristic curves to compare the predictive capability of childhood and adult height instrument scores i n the Avon Longitudinal Study of Children and Parents (ALSPAC). (A) Mean age 9.9 years in the ALSPSAC offspring (B) mean a ge 50.8 years in the ALSPAC mothers. Continuous height in the ALSPAC cohort was dichotomised based on the 50th centile t o separatee above and below average height. AUC=area under curve

Consistent with previous studies (32), we found that adulthood height genetic variants have smaller effect estimates in within-sibship analyses compared to population estimates (shrinkage 10%; 95% CI 6%, 14%). In contrast, we found limited evidence for differential effects of childhood height genetic variants in within-sibship models (shrinkage −2%; 95% CI −7%, 3%). Assortative mating on height is thought to be the primary mechanism underlying the observed within-sibship shrinkage for adulthood height. The lack of evidence for within-sibship shrinkage for childhood height suggests that assortment is typically on adulthood rather than childhood height. Analysing the spouse pairs in UKB supported this hypothesis as the correlation between spouses was markedly higher when comparing their adulthood height measures (r=0.206, P<1×10 ^−300^) in comparison to the correlation of their childhood height (r=0.036, P=1×10 ^−15^).

### Applying lifecourse Mendelian randomization to evaluate whether childhood height has a direct or indirect effect on cardiovascular disease endpoints

Applying univariable MR provided strong evidence that being taller in childhood has a protective effect on risk of later life CAD (OR=0.78 per change in height category, 95% CI=0.70 to 0.86, P=4×10 ^−10^) and stroke (OR=0.93, 95% CI=0.86 to 1.00, P=0.03), but increases risk of later life AF (OR=1.69, 95% CI=1.63 to 1.75, P=3×10 ^−60^) and TAA (OR=1.47, 95% CI=1.34 to 1.60, P=4×10 ^−9^). Limited evidence of an effect was found on risk of PAD (OR=1.02, 95% CI=0.90 to 1.13, P=0.80) (Supplementary Table 4). These effect estimates were typically supported by pleiotropy robust methods with the exception of evidence supporting the effect of childhood height on risk of stroke (Supplementary Table 4) as well as its subtypes (Supplementary Table 5).

In a multivariable setting, evidence of a protective effect between childhood height on CAD and stroke was reduced when accounting for adulthood height in the model (Figure 3). In contrast, there was evidence of a direct effect for childhood height on risk of later life AF (OR=1.61, 95% CI=1.42 to 1.79, P=5×10 ^−7^) and TAA (OR=1.55, 95% CI=1.16 to 1.95, P=0.03) after accounting for adulthood height in the multivariable model (Supplementary Table 6). Evidence of a direct effect on TAA became stronger after excluding instruments located at genetic loci responsible for encoding the fibrillin family of proteins known to play a role in risk of Marfan syndrome (OR=1.62, 95% CI=1.10 to 2.38, P=0.02) (Supplementary Table 7). There was limited evidence that our instruments for childhood and adult height influence natural hair colour as a negative control analysis to investigate potential population stratification in our study (Supplementary Table 8).

**Figure 3:**
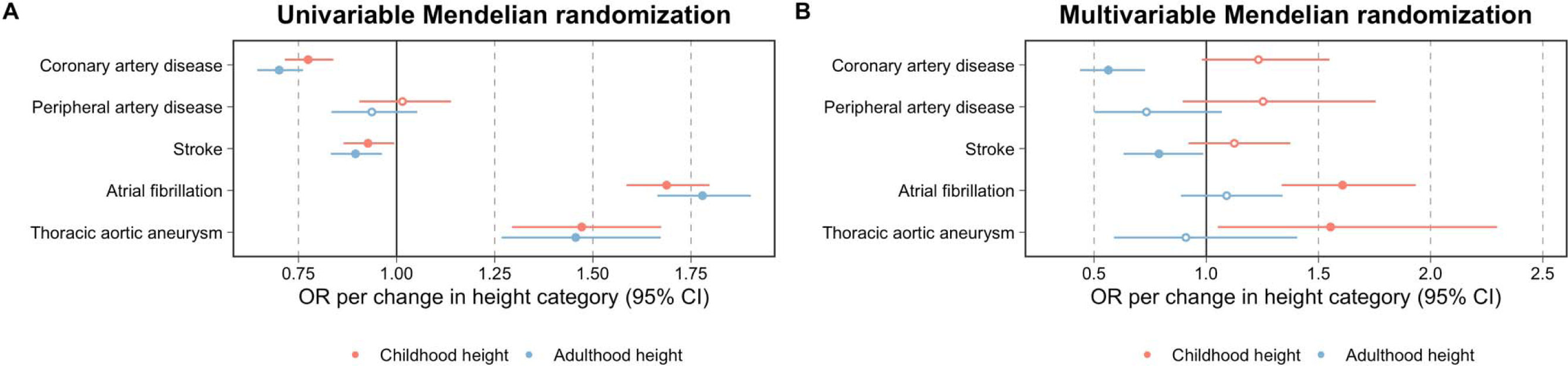
Forest plots portraying thee direct and indirect effects for genetically predicted childhood height (age 10 years) on 5 di fferent disease outcomes. Estimates are displayed as odds ratios (OR) with 95% confidence intervals (95% CI) derived from (A) univariable and (B) multivariable Mendelian randomization analyses.

## Discussion

In this study, we have applied a lifecourse Mendelian randomization approach to evaluate the direct and indirect effects of childhood height on risk of 5 cardiovascular endpoints. Our findings suggest that individuals who are taller in early life typically have lower risk of coronary artery disease and that this is likely due to the causal pathway involving adulthood height. Conversely, childhood height provided evidence of a direct effect on elevated risk of atrial fibrillation and thoracic aortic aneurysm in later life after accounting for the effect of adulthood height.

The aetiological relationship between height and coronary heart disease has been extensively studied with evidence of an inverse relationship being identified by multiple study designs. The exact underlying mechanisms remain unclear although the role of potential intermediate traits such as lower blood pressure and a favourable lipid profile have been postulated. Findings from this study suggest that, although being taller in childhood has a protective effect on risk of coronary heart disease in adulthood, this is likely attributed to the long-term consequence of remaining tall throughout the lifecourse. Similar conclusions were found when analysing stroke as an outcome which likewise corroborates findings from observational studies, albeit with weaker support from pleiotropy robust methods.

In contrast, our findings provide evidence that taller individuals in childhood are at elevated risk of atrial fibrillation and thoracic aortic aneurysm in later life. Evidence of a genetically predicted effect of adulthood height on both of these outcomes has been found by previous MR studies (44). Applying the lifecourse MR approach in this study suggests that these effects may be attributed to a long-term consequence of being taller during early life, highlighting a developmental role as an explanation for these findings. With respect to atrial fibrillation, the effect of thyrotropin on both childhood growth and atrial fibrillation risk has been proposed as a mechanism (45). Regarding thoracic aortic aneurysm, fibrillin mutations have been shown to relate both to greater childhood height and aortic root diameter (46). We show that excluding the fibrillin genic regions does not remove the childhood height – thoracic aortic aneurysm causal link, suggesting that greater height achieved through additional mechanisms also influences later disease risk.

Notably, the inferences made in this work would have been extremely challenging to make without the use of human genetic data, particularly given the strong degree of correlation between childhood and adult height of the UK Biobank participants. Validation of the genetic scores derived for these measures suggested that the genetically predicted effects of height at these timepoints in the lifecourse (i.e. at age 10 and mean age 55 years) are more challenging to separate compared to the scores we previously derived for body size at these same timepoints. However, the use of reported childhood height data from the UK Biobank allowed us to analyse a huge sample size (n=454,023) compared to previous endeavours (n=18,737) (30) and we found that our genetic estimates are strongly correlated with those of measured childhood height (rG=0.90, 95% CI=0.82-0.98). Future work in this space could focus on other timepoints across the lifecourse to try and separate more granular lifestage specific effects of height as and when relevant datasets become available. Tissue specific expression or DNA methylation data could also contribute to elucidating the mechanisms of disease risk (47).

### Conclusions

Our study suggests that the association between childhood height and reduced risk of coronary artery disease and stroke is likely attributed to the causal pathway involving adulthood height (i.e. taller children on average grow to become tall adults). In contrast, our results indicate that childhood height may increase later life risk of atrial fibrillation and thoracic aortic aneurysm independently of adult height. This highlights a need for further in-depth research into the developmental origins of these disease outcomes to gain insight into the critical windows by which early life growth exerts influences on their lifelong risk.

## Data Availability

All summary-level data analysed in this study can be obtained from the sources referenced within. ALSPAC data can be accessed by submitting a research proposal at https://www.bristol.ac.uk/alspac/researchers/access/

## Acknowledgements

We would like to thank the authors of all the GWAS who made their summary statistics available for the benefit of this work and participants of the UK Biobank study. We want to acknowledge the participants and investigators of the FinnGen and 1958NCDS studies. We are extremely grateful to all the families who took part in this study, the midwives for their help in recruiting them and the whole ALSPAC team, which includes interviewers, computer and laboratory technicians, clerical workers, research scientists, volunteers, managers, receptionists and nurses. This work was supported by the Integrative Epidemiology Unit which receives funding from the UK Medical Research Council and the University of Bristol (MC_UU_00011/1). GDS conducts research at the NIHR Biomedical Research Centre at the University Hospitals Bristol NHS Foundation Trust and the University of Bristol. The views expressed in this publication are those of the author(s) and not necessarily those of the NHS, the National Institute for Health Research or the Department of Health. The authors would like to acknowledge the use of the University of Exeter High-Performance Computing (HPC) facility in carrying out this work (MRC Clinical Research Infrastructure award MR/M008924/1). The study was supported by the National Institute for Health and Care Research Exeter Biomedical Research Centre. The views expressed are those of the author(s) and not necessarily those of the NIHR or the Department of Health and Social Care.

